# Discovery and Validation of SVEP1 and Other Novel Cardiovascular Biomarkers For Patients with Kidney Failure On Maintenance Hemodialysis

**DOI:** 10.64898/2026.04.23.26348442

**Authors:** Yue Ren, Tariq Shafi, Mark R. Segal, Hongzhe Li, Alexander R. Pico, Min-Gyoung Shin, Jeffrey R. Schelling, John D. Hulleman, Jiang He, Changwei Li, Hernan Rincon Choles, Julia Brown, Mirela Dobre, Rupal Mehta, Rajat Deo, Anand Srivastava, Jonathan Taliercio, Stephen M. Sozio, Bernard Jaar, Michelle M. Estrella, Wei Chen, Glenn M. Chertow, Rulan Parekh, Peter Ganz, Ruth F. Dubin, the CRIC Study Investigators

## Abstract

**Background:** Patients with kidney failure undergoing maintenance hemodialysis suffer high rates of major adverse cardiovascular events(MACE) that are not accurately predicted by traditional cardiovascular risk models. There is an urgent need to identify novel, modifiable cardiovascular risk factors for these patients.

**Methods:** We analyzed associations of 6287 circulating proteins with MACE among 1048 participants undergoing hemodialysis in the Chronic Renal Insufficiency Cohort(CRIC) (14-year follow-up) with validation in the Predictors of Arrhythmic and Cardiovascular Risk in End-Stage Renal Disease study(PACE) (7-year follow-up). In both cohorts, proteins were measured shortly after dialysis initiation and one year later. We compared protein-based risk models derived by elastic net regression to the Pooled Cohort Equations(PCE) optimized for these cohorts(Refit PCE), and to an Expanded Refit PCE that included Troponin T and N-terminal pro-B-type natriuretic peptide.

**Results:** In CRIC, 149 proteins were associated with MACE at false discovery rate<0.05. Among 22 proteins significant at Bonferroni p<8×10^-6^, proteins that validated in PACE included Sushi von Willebrand factor type A EGF and pentraxin domain-containing protein 1(SVEP1), Complement component C7, R-spondin 4, Tenascin, Fibulin-3 and Fibulin-5. Complement pathways were prominent in network analyses. SVEP1 surpassed other markers by statistical significance, with CRIC HR per log_2_ 1.8 (p=2.1×10^-12^) and HR per annual doubling 1.6 (p=6.8×10^-6^). For 2-year MACE, AUC(95%CI) for SVEP1 alone was 0.72(0.59, 0.84) in CRIC, and 0.73(0.63, 0.81) in PACE. SVEP1 surpassed the Expanded Refit PCE in CRIC (0.61 (0.48, 0.73)) (p=0.038). In the pooled CRIC + PACE cohort, SVEP1 AUC(95%CI) (0.79(0.70, 0.88)) surpassed Refit PCE (0.61(0.51, 0.72)) (p=0.004).

**Conclusions:** SVEP1, a 390 kDa protein unlikely to be renally cleared, surpassed over 6000 other proteins and by itself outperformed traditional clinical risk models in predicting MACE in two populations of patients undergoing maintenance hemodialysis. Future studies should provide mechanistic insights behind these findings.

**Key Points:**

1. Patients with kidney failure undergoing hemodialysis have 20-fold higher cardiovascular mortality compared to the general population, and conventional risk factors have low prognostic utility for these patients.
2. By applying large-scale circulating proteomics in two independent hemodialysis cohorts, we have discovered >20 novel proteins that predict major adverse cardiovascular events(MACE).
3. Sushi von Willebrand factor type A EGF and pentraxin domain-containing protein 1(SVEP1) surpassed >6000 individual proteins and clinical factors for predicting MACE.

## INTRODUCTION

For the global population of ∼3 million patients with kidney failure undergoing maintenance hemodialysis,^1,2^ cardiovascular disease(CVD) mortality is 20-fold higher than that of the general population.^3^ CVD risk factors observed in populations with normal or near normal kidney function, including hypercholesterolemia and obesity, exhibit paradoxical associations with CVD outcomes in the kidney failure population;^4^ indeed, randomized placebo controlled trials of statins showed no benefit.^5,6^ The urgent need to identify novel, modifiable CVD risk factors for the kidney failure population was highlighted in a statement by Kidney Disease Improving Global Outcomes, who called for the “development and validation of end-stage renal disease-specific CVD risk prediction scores.”^7^

Accordingly, to identify novel risk factors for CVD in patients with kidney failure receiving maintenance hemodialysis that are biologically based, we conducted a large-scale proteomics investigation of cryopreserved plasma samples from participants undergoing maintenance hemodialysis enrolled in the Chronic Renal Insufficiency Cohort (CRIC) study, with 14 years of follow-up for adjudicated major adverse cardiovascular outcomes (MACE). We validated our findings in Predictors of Arrhythmic and Cardiovascular Risk in End-Stage Renal Disease (PACE) study. In numerous disease settings, multi-protein risk models predict clinical outcomes better than traditional clinical models^8,9^ or genetic risk scores.^10^ Furthermore, unlike genetic risk scores or many clinical risk factors, protein levels are mutable, reflecting disease activity and medication changes, suggesting that assessing proteomic cardiovascular risk at more than one time point may yield additional prognostic information.^11^ We thus measured 6287 plasma proteins with SomaScan V4.1 in both cohorts at two time points, corresponding to incident and prevalent time period following dialysis initiation.^12,13^ Our aims were 1) to identify novel proteins with higher prognostic utility for MACE compared to conventional risk markers 2) to identify dynamic factors whose change between the two time points predict MACE; 3) to identify biological pathways associated with MACE; and 4) to construct protein-based risk models for MACE and compare their performance to conventional risk models.

## BRIEF METHODS

### CRIC and PACE Participants

Between 2004 and 2020, 1123 CRIC^14^ participants reached kidney failure and started dialysis. Among them, 251 had no further study visits and another 232 provided no blood samples after starting dialysis. We included 630 CRIC participants with kidney failure receiving maintenance hemodialysis, enrolled from 13 sites at 16 different study visits **(Supplement 1).** We ran SomaScan V4.1 on plasma EDTA samples drawn at the first study visit following initiation of dialysis (incident period); we assayed samples on 421 participants who remained on hemodialysis a mean of 1.1 ± 0.8 years following the first proteomics sample (prevalent period). External validation was performed using samples from the first two of four annual study visits of PACE,^15^ a study of participants recently initiated on hemodialysis, enrolled between 2008-2012 from 27 dialysis units in the Baltimore area.

### SomaScan V4.1

The SomaScan V4.1 menu includes 7596 aptamers. We excluded 308 aptamers paired with non-human proteins, and 117 incompletely characterized investigational aptamers. We excluded 0.4% of aptamers (32 for CRIC, 31 for PACE) that saturated the SomaScan assay due to high concentrations, and 4 proteins with intra-assay CV>50% in duplicate samples run in this study. Given that some proteins are measured by two or more aptamers, we analyzed 7135 aptamers (6287 unique proteins) in CRIC, and 7140 aptamers (6291 unique proteins) in PACE. We have previously published quality control analyses of SomaScan in patients with kidney failure.^16^ Aptamers for SVEP1 and numerous other proteins have been validated by mass spectrometry.^17^ (SomaScan V4.1 is further described in **Supplement 2 and 3**.)

### Study Outcomes

In CRIC, cardiovascular events were adjudicated by two physicians after reviewing imaging, laboratory data, hospital notes and death certificates, and categorized as probable or definite myocardial infarction(MI), heart failure(HF), stroke or CVD death, as previously described.^14,18^ CVD and non-CVD deaths were ascertained from next of kin, death certificates, obituaries, hospital records, the Social Security Death Master File, and the National Death Index. There was a small proportion of deaths labelled as “unknown cause” and given the high frequency of sudden cardiac arrests in this population,^19^ we included these as CVD deaths in the composite outcome of MACE. Hereafter, “CVD deaths” refers to events labelled CVD death or unknown death in either cohort. The composite CVD outcome of MACE consisted of time to the first of probable or definite MI, HF, stroke, or CVD death. Censoring for kidney transplant or change to peritoneal dialysis is described in **Supplemental Methods**.

### Statistical Analysis

Our primary approach was to consider CRIC as the discovery cohort and PACE the validation cohort. We used Cox proportional hazards regression and alternatively, accounted for competing risk of non-CVD death using the Fine Gray method.^20^ We aimed to evaluate novel protein CVD risk factors in the context of conventional CVD risk factors in the Pooled Cohort Equations(PCE):^21^ age, sex, race, total cholesterol, high-density lipoprotein cholesterol, systolic blood pressure, hypertension treatment, diabetes, current smoking. PCE variables are readily applied to patients with kidney failure, in whom eGFR and proteinuria (variables included in Predicting Risk of CVD EVENTs(PREVENT) equations^22^) are often not available. Additionally, we enhanced the PCE model by adding two established cardiovascular biomarkers in the kidney failure population, Troponin T(TnT) and N-terminal pro-B-type natriuretic peptide (NT-proBNP.)^23^ We created three conventional models: Original PCE,^21^ and a Refit PCE that included sex and self-reported race, with the coefficients of all PCE variables refit to the training set to optimize its performance in the kidney failure population. The third clinical risk model, the Expanded Refit PCE, was devised by refitting PCE variables, plus aptamer measures of TnT and NT-proBNP. We previously have shown good correlation between traditional and aptamer assays for TnT (*rho* 0.68) and NT-proBNP(*rho* 0.80) in plasma of patients with kidney failure.^16^ Aiming to develop multi-protein and hybrid clinical-protein models, we applied elastic net regression in these datasets: 1) 6287 proteins; 2) 6287 proteins + PCE variables; 3) in the pooled cohort of CRIC and PACE (see below), we additionally evaluated 6287 proteins, PCE variables, and dialysis-related factors (time since dialysis initiation, diastolic blood pressure, body mass index, triglycerides, albumin, hemoglobin, calcium, phosphorus, parathyroid hormone(PTH)). Risk models were derived in a training set obtained as a random partition of 80% of the CRIC participants, tested in the remaining 20%, then validated in the full PACE cohort. After finding that SVEP1 surpassed other markers in both cohorts, we further examined SVEP1 in a pooled analysis of CRIC and PACE which yielded a larger sample size suitable for subgroup analyses and risk model comparisons. Subgroup analyses were thus performed in the full pooled cohort; risk models were trained and then tested in pooled cohort 80% and 20% sets, respectively.

### External Validation

We validated the 22 proteins associated with MACE outcome in CRIC at p<8×10-6, in PACE, in full cohort and full follow-up time, designating validation as FDR<0.05. Risk models, either with published coefficients (as for PCE), or with coefficients refit to the CRIC 80% testing set, (Refit PCE or Expanded Refit PCE) were also validated in the full PACE cohort.

Additional information about clinical covariates, outcome adjudication, functional enrichment and elastic net regression is found in **Supplemental Methods.**

## RESULTS

### Characteristics of CRIC and PACE Participants

CRIC and PACE participants had similar proportion of women (CRIC 40%, PACE 41%), Black race (CRIC 57%, PACE 71%), and participants with kidney failure due to diabetes (CRIC 44%, PACE 36%). Compared to PACE, CRIC participants were older, and more likely to have a history of MI, HF or stroke. PACE participants were more likely to have a tunneled catheter as opposed to permanent vascular access (arteriovenous fistula or graft), higher PTH, and lower calcium. (**Table 1**)

**Table 1.** Baseline Characteristics for Participants in CRIC and PACE.

| Characteristic<br>N(%) or Median[IQR] |  | CRIC<br>N=630 | PACE<br>N=418 |
| --- | --- | --- | --- |
| Age (years) |  | 62 [54, 70] | 55 [47, 64] |
| Females |  | 254 (40) | 171 (41) |
| Race | White | 181 (29) | 114 (27) |
|  | Black | 358 (57) | 296 (71) |
|  | Other | 91 (14) | 8 (2) |
| History of myocardial infarction |  | 249 (40) | 36 (9) |
| History of heart failure |  | 190 (30) | 84 (20) |
| History of stroke |  | 120 (19) | 25 (6) |
| History of diabetes |  | 466 (74) | 244 (58) |
| Cause of kidney failure | Diabetes | 278 (44) | 149 (36) |
|  | Hypertension | 107 (17) | 110 (26) |
|  | Glomerulonephritis | N/A | 54 (13) |
|  | Other/unknown | 245 (39) | 103 (26) |
| Months since starting dialysis |  | 7.4 [4, 12] | 3.8 [3, 6] |
| Vascular access for hemodialysis |  | 376 (70) | 142 (34) |
| Three dialysis sessions per week |  | 591 (97) | 414 (100) |
| Current smoking |  | 73 (12) | 142 (34) |
| Systolic blood pressure (mmHg) |  | 127 [113, 144] | 136 [119, 152] |
| Diastolic blood pressure (mmHg) |  | 64 [56,73] | 73 [64, 82] |
| Body mass index (kg/m<sup>2</sup>) |  | 30 [26, 35] | 28 [24, 34] |
| Total cholesterol (mg/dl) |  | 152 [128, 183] | 158 [136, 189] |
| Triglycerides (mg/dl) |  | 126 [88, 183] | 117 [87, 157] |
| LDL-C (mg/dl) |  | 77 [58, 106] | 81 [60, 107] |
| HDL-C (mg/dl) |  | 42 [34, 52] | 50 [40, 63] |
| Hemoglobin (g/dL) |  | 12 [11,13] | 11 [10,11] |
| Calcium (mg/dL) |  | 9.2 [8.7, 9.6] | 8.6 [8.2, 8.9] |
| Phosphorus (mg/dL) |  | 4.4 [3.9, 5.0] | 4.9 [4.3, 5.6] |
| Parathyroid hormone (pg/ml) |  | 198 [130, 302] | 428 [273, 620] |
| Albumin (g/dL) |  | 3.8 [3.5,4.1] | 3.6 [3.3,3.9] |
| Table includes self-reported sex and self-reported race in PACE and CRIC; these data were not collected from United States Renal Data System |  |  |  |

### MACE Outcomes in CRIC and PACE

Over 5 years starting from the first proteomic time point, 266 (42%) CRIC participants compared to 146 (35%) of PACE participants experienced MACE. Five-year rates of non-fatal MI, HF, and stroke were similar in CRIC and PACE, while a larger proportion of CRIC participants experienced CVD deaths (13%) vs PACE (8%). (**Table 2**; censoring and detailed description of events are found in **Supplement 4)**

**Table 2.** Events in CRIC and PACE.

| Event | CRIC Follow-up:<br>Maximum 14 years<br>Median [IQR] 2.9[1.0, 5.2] |  | PACE Follow-up:<br>Maximum 7 years<br>Median [IQR] 1.6[0.47, 2.8] |  |
| --- | --- | --- | --- | --- |
|  | 5-year | All follow-up | 5-year | All follow-up |
| **First MACE event (N, %)** |  |  |  |  |
| Myocardial infarction | 44 (7) | 51 (8) | 24 (6) | 26 (6) |
| Heart Failure | 115 (18) | 131 (21) | 72 (17) | 73 (18) |
| Stroke | 13 (2) | 19 (3) | 16 (4) | 16 (4) |
| CVD death | 77(13) | 107(18) | 32 (8) | 38 (9) |
| **MACE outcome (N, %)** | 266 (42) | 327 (52) | 146 (35) | 153 (37) |
| **Other Events (N,%)** |  |  |  |  |
| Non-CVD deaths | 45 (7) | 53 (8) | 76 (18) | 85 (20) |
| Transplant | 77 (12) | 88 (14) | 56 (13) | 59 (14) |
| Lost to follow-up due to study discontinuation or change to peritoneal dialysis | 71 (11) | 157 (25) | 28 (7) | 28 (7) |
| **Total all-cause mortality (N,%)** | 222(35) | 328(52) | 170(40) | 199(48) |

### Associations of Individual Proteins with MACE

In CRIC, we observed higher numbers of proteins significantly associated with MACE for samples measured at the second time point compared to the first time point (149 proteins vs. 53 proteins, significant at false discovery rate (FDR)<0.05) despite fewer participants and events subsequent to time point 2. This finding motivated us to prioritize time point 2, when virtually all patients had been undergoing hemodialysis for ≥ 1 year and could be considered ‘prevalent’ patients as opposed to ‘incident’ patients. Here we report results for proteins assayed at time point 2, and change in protein between time point 1 and 2. Proteins measured at time point 1 (incident time period) and their associations with MACE are shown in **Supplements 5 and 6.**

For CRIC prevalent participants (time point 2 samples), there were 22 proteins significant at Bonferroni significance threshold of p<8×10^-6^. We display 12 of these 22 proteins that successfully validated in PACE in **Table 3**. Sushi von Willebrand factor type A EGF and pentraxin domain-containing protein 1 (SVEP1), had the most robust level of statistical significance across all the models, with HR=2.1 per log_2_, p=2.0×10^-12^ in unadjusted analyses, and 1.8 per log_2_, p=1.3×10^-7^ after accounting for competing risk of non-CVD death and adjusting for traditional and dialysis-related cardiovascular risk factors. All twelve proteins remained significant at p<0.05 after multivariable adjustment in Cox or Fine-Gray models. For each of these 12 top proteins, the association is largely linear (**Figure 1**).

**Figure 1.**
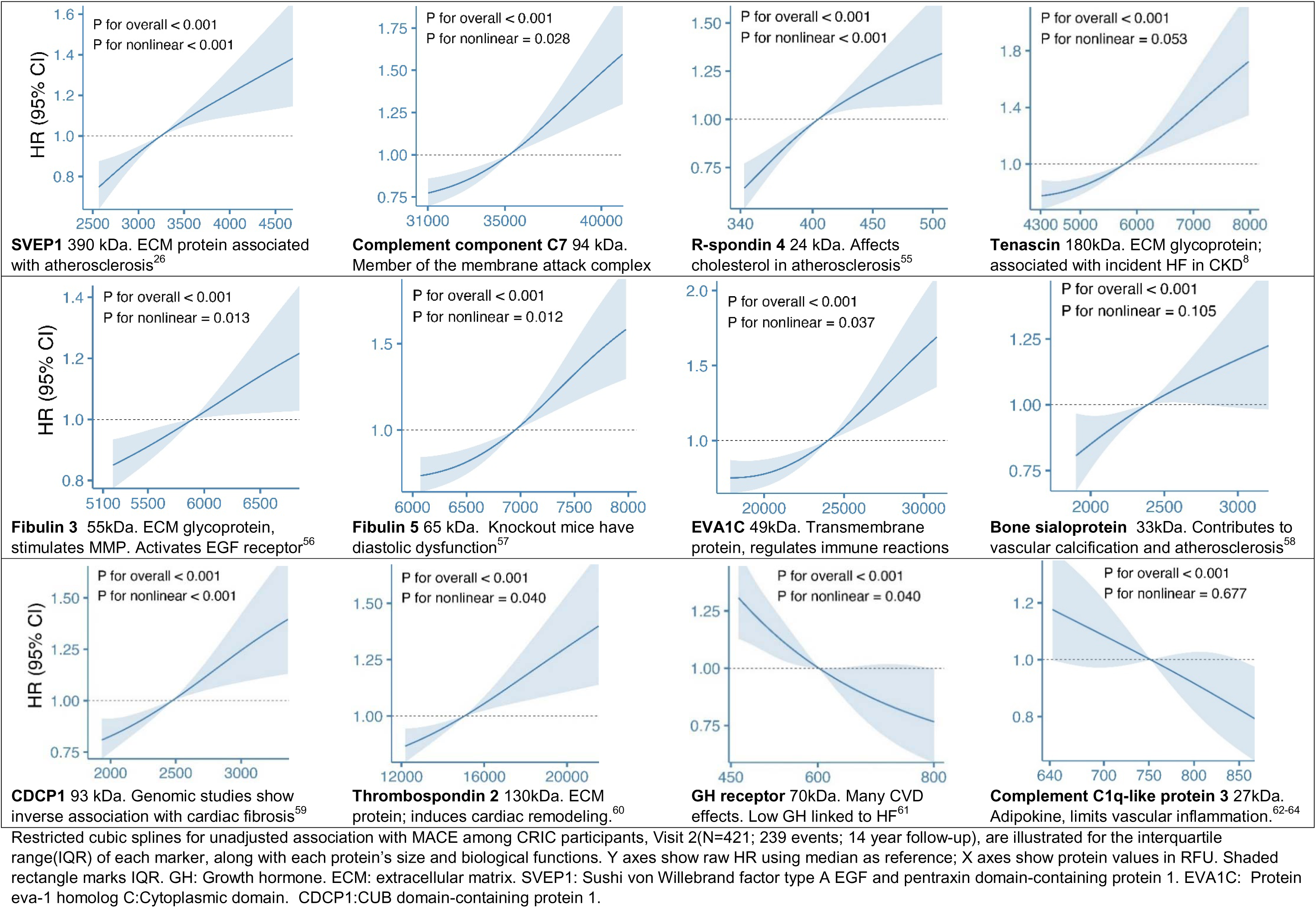
Distribution, Splines and Main Biology for Top Proteins Associated with MACE. Restricted cubic splines for unadjusted association with MACE among CRIC participants, Visit 2(N=421; 239 events; 14 year follow-up), are illustrated for the interquartile range(IQR) of each marker, along with each protein’s size and biological functions. Y axes show raw HR using median as reference; X axes show protein values in RFU. Shaded rectangle marks IQR. GH: Growth hormone. ECM: extracellular matrix. SVEP1: Sushi von Willebrand factor type A EGF and pentraxin domain-containing protein 1. EVA1C: Protein eva-1 homolog C:Cytoplasmic domain. CDCP1:CUB domain-containing protein 1.

**Table 3.** Proteins Associated with MACE In CRIC and PACE.

|  | No adjustment for covariates |  | Adjusted for PCE variables |  |  |  | Adjusted for PCE + Dialysis-related factors |  |  |  |
| --- | --- | --- | --- | --- | --- | --- | --- | --- | --- | --- |
| **CRIC N=421** | No competing risk |  | No competing risk |  | Non-CVD death treated as competing risk |  | No competing risk |  | Non-CVD death treated as competing risk |  |
|  | HR | p | HR | p | HR | p | HR | p | HR | p |
| **Complement component C7** | 3.20 | 8.7x10<sup>-9</sup> | 3.07 | 6.4x10<sup>-7</sup> | 2.68 | 1.5x10<sup>-6</sup> | 2.8 | 1.5x10<sup>-5</sup> | 2.47 | 3.3x10<sup>-5</sup> |
| **R-spondin 4** | 2.82 | 6.9x10<sup>-11</sup> | 2.55 | 9.8x10<sup>-7</sup> | 1.96 | 3.1x10<sup>-5</sup> | 2.4 | 4.2x10<sup>-5</sup> | 1.9 | 0.0019 |
| **Fibulin-5** | 2.74 | 2.5x10<sup>-6</sup> | 2.19 | 0.0011 | 2.04 | 0.0011 | 2.2 | 0.0019 | 2.0 | 0.0024 |
| **Fibulin 3** | 2.30 | 6.7x10<sup>-6</sup> | 2.01 | 8.5x10<sup>-4</sup> | 1.85 | 7.6x10<sup>-4</sup> | 2.0 | 0.0017 | 1.8 | 0.0048 |
| **SVEP1** | 2.10 | 2.0x10<sup>-12</sup> | 2.17 | 2.0x10<sup>-9</sup> | 1.88 | 8.0x10<sup>-9</sup> | 2.2 | 1.0x10<sup>-8</sup> | 1.9 | 1.3x10<sup>-7</sup> |
| **Tenascin** | 2.05 | 5.5x10<sup>-11</sup> | 1.82 | 3.0x10<sup>-9</sup> | 1.61 | 1.5x10<sup>-8</sup> | 1.8 | 2.7x10<sup>-8</sup> | 1.6 | 2.9x10<sup>-7</sup> |
| **Protein eva-1 homolog C: Cytoplasmic domain** | 1.90 | 2.2x10<sup>-7</sup> | 2.21 | 7.9x10<sup>-8</sup> | 1.85 | 7.0x10<sup>-6</sup> | 2.2 | 3.9x10<sup>-7</sup> | 1.9 | 2.6x10<sup>-5</sup> |
| **Bone sialoprotein** | 1.76 | 4.5x10<sup>-6</sup> | 1.71 | 4.8x10<sup>-5</sup> | 1.54 | 4.3x10<sup>-4</sup> | 1.7 | 0.00011 | 1.6 | 0.00063 |
| **CUB domain-containing protein 1** | 1.68 | 4.5x10<sup>-6</sup> | 1.39 | 0.0085 | 1.38 | 0.0089 | 1.3 | 0.041 | 1.3 | 0.044 |
| **Thrombospondin 2** | 1.60 | 3.4x10<sup>-6</sup> | 1.58 | 8.4x10<sup>-5</sup> | 1.46 | 0.0013 | 1.6 | 0.00014 | 1.4 | 0.0037 |
| **Growth hormone receptor** | 0.52 | 6.2x10<sup>-7</sup> | 0.45 | 1.6x10<sup>-6</sup> | 0.53 | 6.9x10<sup>-6</sup> | 0.47 | 4.0x10<sup>-5</sup> | 0.52 | 5.5x10<sup>-5</sup> |
| **Complement C1q-like protein 3** | 0.38 | 4.9x10<sup>-6</sup> | 0.37 | 2.1x10<sup>-5</sup> | 0.50 | 2.2x10<sup>-4</sup> | 0.32 | 2.6x10<sup>-6</sup> | 0.47 | 5.3x10<sup>-5</sup> |

Proteins shown have HR per log<sub>2</sub> significant at p<8x10<sup>-6</sup> in N=421 CRIC Visit 2 participants, before adjustment for competing risk, using all follow-up time and are also significant at FDR<0.05 in PACE participants. HR for the composite CVD outcome are per log<sub>2</sub> of protein in RFU, evaluated over all follow-up time. Pooled cohort equation (PCE) variables are age, race, sex, log(total cholesterol), log (HDL), log(SBP), hypertension treatment, DM, smoking. Dialysis-related factors are log(months since dialysis initiation), log(DBP), log(BMI), log(triglycerides), serum albumin, hemoglobin, log(calcium), log(phosphate), log(PTH)

### Within-subject Longitudinal Changes in Proteins

Trajectories of change for several novel proteins were associated with MACE in CRIC, with trajectories reported here as annualized percent change and associations as HR per 25% annualized change. In CRIC, SVEP1 median[IQR] change was 2.0% [-18, 29] and HR(95%CI) was 1.12 (1.1, 1.2) (p=7×10^-6^). Chitinase-3-like protein 1, also known as YKL-40, an inflammatory mediator that has been proposed as a biomarker for atherosclerotic CVD and CKD,^24^ had median[IQR] change -1.2% [-17, 19] and HR(95%CI) 1.2 (1.1, 1.3) (p=8.4×10^-6^). Increases in R-spondin 4 or tenascin predicted MACE at p<0.005. Change of numerous proteins had similar associations with MACE in CRIC and PACE, based on a nominal p<0.05 for both cohorts, including markers of cardiac fibrosis, such as calsyntenin-2;^25^ atherosclerosis, such as SVEP1;^26^ natriuretic peptides; and inflammation, such as complement component C1q receptor **(Supplements 12-14).**

### Network Analyses and Functional Enrichment

To understand the biological processes underlying the high risk of MACE in patients with kidney failure on hemodialysis, we performed network analysis of 62 proteins associated with MACE at FDR<0.01 from CRIC time point 2, in Search Tool for the Retrieval of Interacting Genes/Proteins(STRING). The largest network that was returned included 64 proteins and was enriched with proteins in the complement pathways, insulin-like growth factor binding proteins, fibulins, and natriuretic peptides. (**Figure 2**) Over-representation analyses also demonstrated high significance for the Complement System term in Wikipathways (p<10^-17^). **(Supplement 15)**.

**Figure 2.**
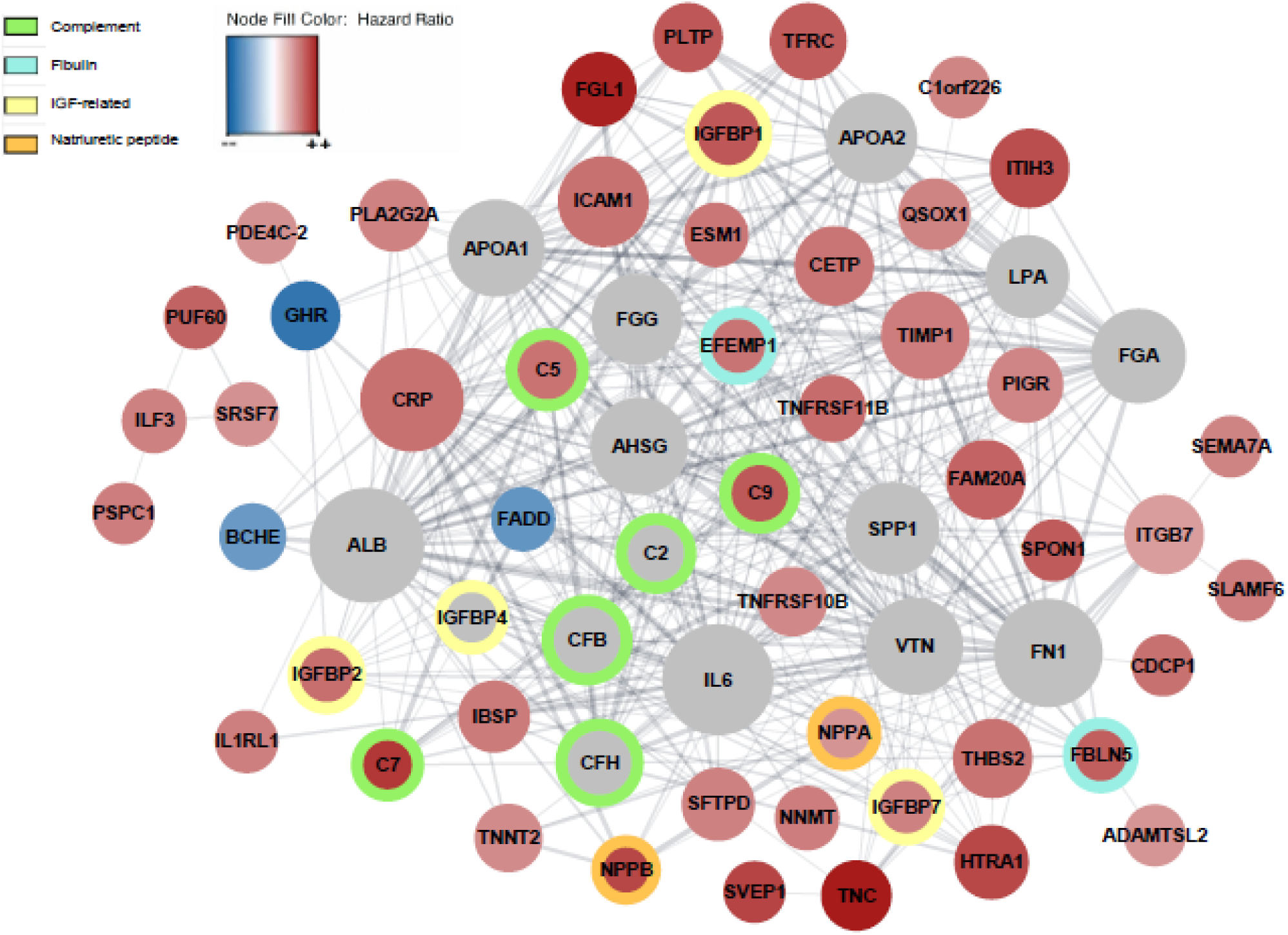
Biological Networks Associated with MACE. **Legend:** In the STRING network, red or blue nodes correspond to HR>1 or HR<1, and node shading corresponds to transformed HR = (-(1/HR)). If the protein was not among top proteins associated with the composite CVD outcome, there is no corresponding HR value and it is shaded in grey. Node size reflects the degree, or number of connections between the protein node and other proteins. Protein names are listed in Supplement 13.

### Proteomic Risk Models Compared to Conventional Risk Models in CRIC and PACE

When applied to protein datasets alone, elastic net regression selected 1 to 5 proteins, the number depending on tuning parameters; each version included SVEP1, and the highest AUC was achieved in models with SVEP1 alone and no other protein. When elastic net was applied to proteins + PCE variables, no PCE variables were selected, and the best model was SVEP1 alone **(Supplement 16).** For the 2-year time horizon in CRIC, SVEP1 AUC(95%CI) was 0.717 (0.59, 0.84), higher than either the Original PCE(0.572 (0.41, 0.72)) (p=0.036) or Expanded Refit PCE(0.609 (0.48, 0.73)) (p=0.038)). In PACE, AUC(95%CI) for SVEP1 (with the same coefficient as in CRIC) was 0.732 (0.64, 0.81) (**Figure 3**). Time-dependent AUCs demonstrate higher AUC for SVEP1 than the Expanded Refit PCE for 2-year follow-up in CRIC. SVEP1 surpassed the Expanded Refit PCE in the PACE validation cohort during years 2-7. (**Figure 4**; specific p-values listed in **Supplemental Fig. 1.)**

**Figure 3.**
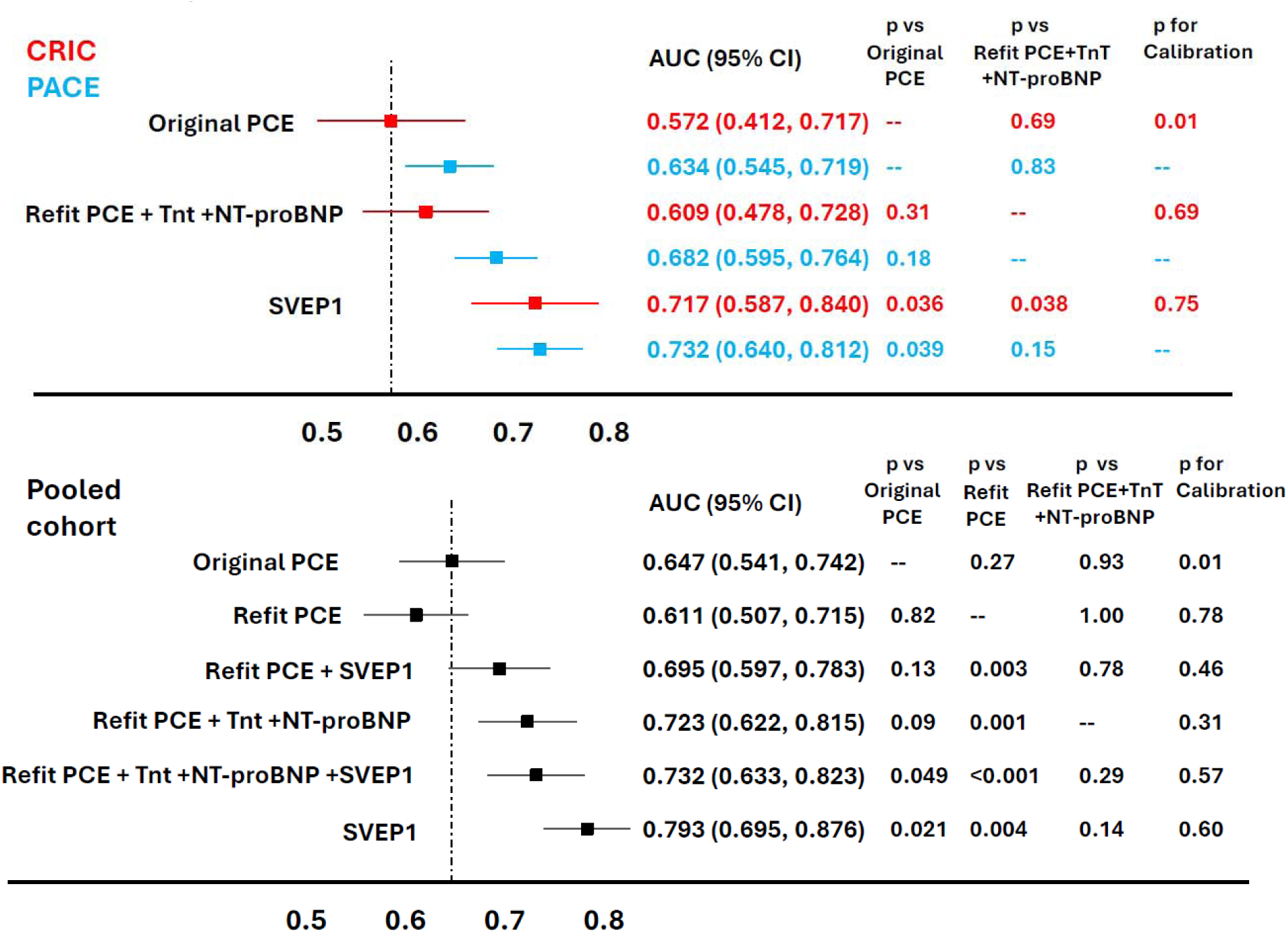
Conventional Risk Models vs. SVEP1 for Two-Year MACE. **Legend**: Discrimination (point estimates and standard errors for AUC) and calibration for risk models predicting time to the composite CVD outcome occurring within 2 years are shown for participants who have undergone hemodialysis for _≥_ 1year (prevalent time period). In the top panel, protein selection and model fitting was performed in CRIC 80% training set, and AUCs were evaluated in the CRIC 20% set (84 participants, 27 events) and then validated in PACE full cohort (226 participants, 48 events). For the pooled cohort, AUCs are shown for the 20% testing set (129 participants, 32 events). One-sided p-values were obtained with paired bootstrapping t-tests. PCE: Pooled Cohort Equations. TnT: Troponin T. NT-proBNP: N-terminal pro-B-type natriuretic peptide. SVEP1: Sushi von Willebrand factor type A EGF and pentraxin domain-containing protein 1.

**Figure 4.**
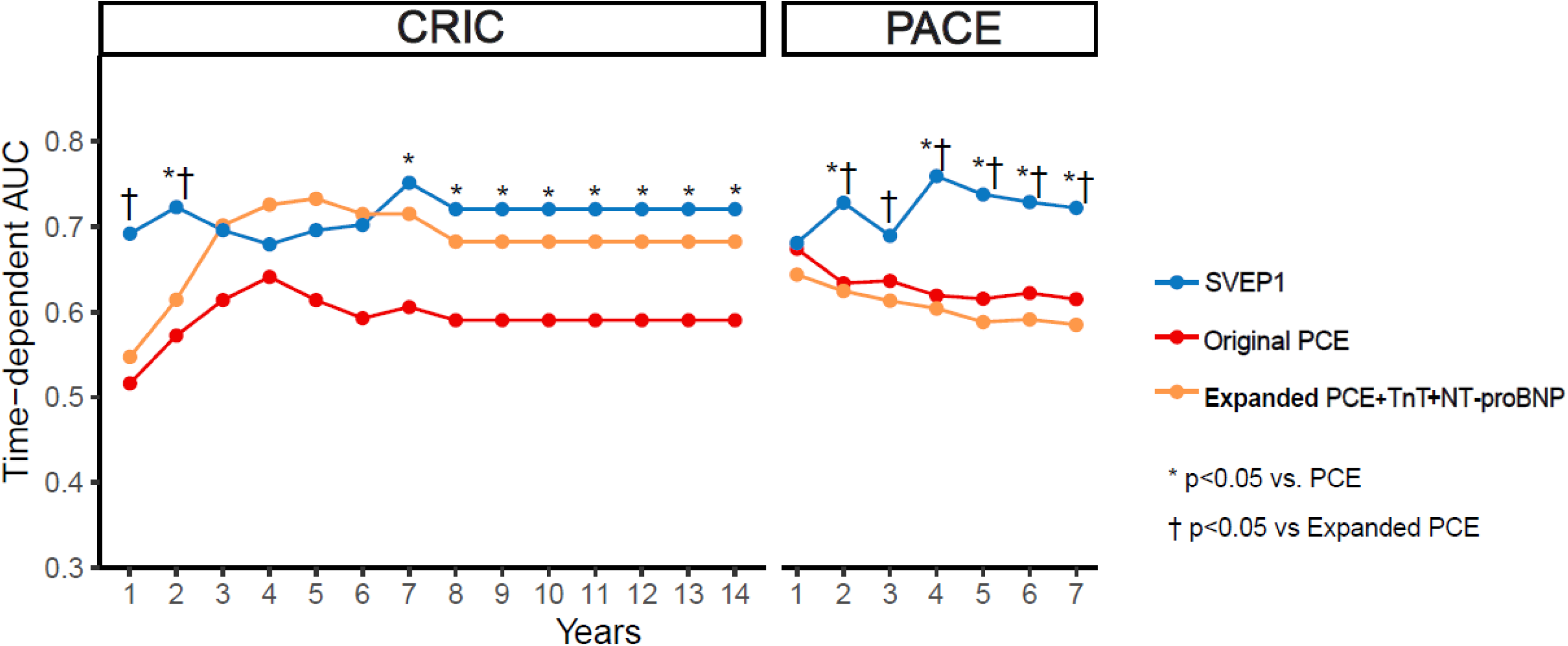
Time-dependent Area Under the Curve for Conventional Models and SVEP1. **Legend:** Time-dependent Areas Under the Receiver Operating Characteristic curves (AUCs) for MACE are shown for each risk model or protein using Visit 2, prevalent period. Expanded PCE + TnT + NT-proBNP and the model comprised of SVEP1 alone were fit to CRIC 80% training set. AUCs are shown for each model in CRIC 20% testing set (N=84) and PACE full cohort(N=226). PCE: Pooled Cohort Equations. TnT: Troponin T. NT-proBNP: N-terminal pro-B-type natriuretic peptide. SVEP1: Sushi von Willebrand factor type A EGF and pentraxin domain-containing protein 1.

### SVEP1 Compared to Conventional Models and Single Markers in the Pooled Cohort

While the high rate of the CVD events lent sufficient statistical power to analyses in separate cohorts, we nevertheless recognized that risk modeling, including any subgroup analyses, alternate time horizons, and analyses of individual events within MACE, would be facilitated by a larger sample size in a pooled cohort of CRIC + PACE. In the pooled cohort, protein models and refit conventional models were trained in 80% of the pooled CRIC+PACE cohort, and then the AUC(95%CI) performance was obtained in the 20% testing set. AUC(95%CI) for SVEP1 was 0.793 (0.70, 0.88), significantly better than the Original (0.647 (0.54, 0.74)) (p=0.02) or Refit PCE (0.611 (0.51, 0.72)) (p=0.004). Adding SVEP1 to the Refit PCE significantly improved AUC(95%CI) from 0.611(0.51, 0.72) to 0.695 (0.60, 0.78) (p=0.003). (**Figure 3; Supplement 17, 18).** We compared AUCs for individual protein biomarkers TnT, NT-proBNP and SVEP1 using the pooled cohort, and observed the highest AUC(95%CI) for SVEP1 (0.793 (0.70, 0.88)), compared to NT-proBNP (0.722 (0.63, 0.82)) (p=0.05). (**Figure 5**)

**Fig. 5:**
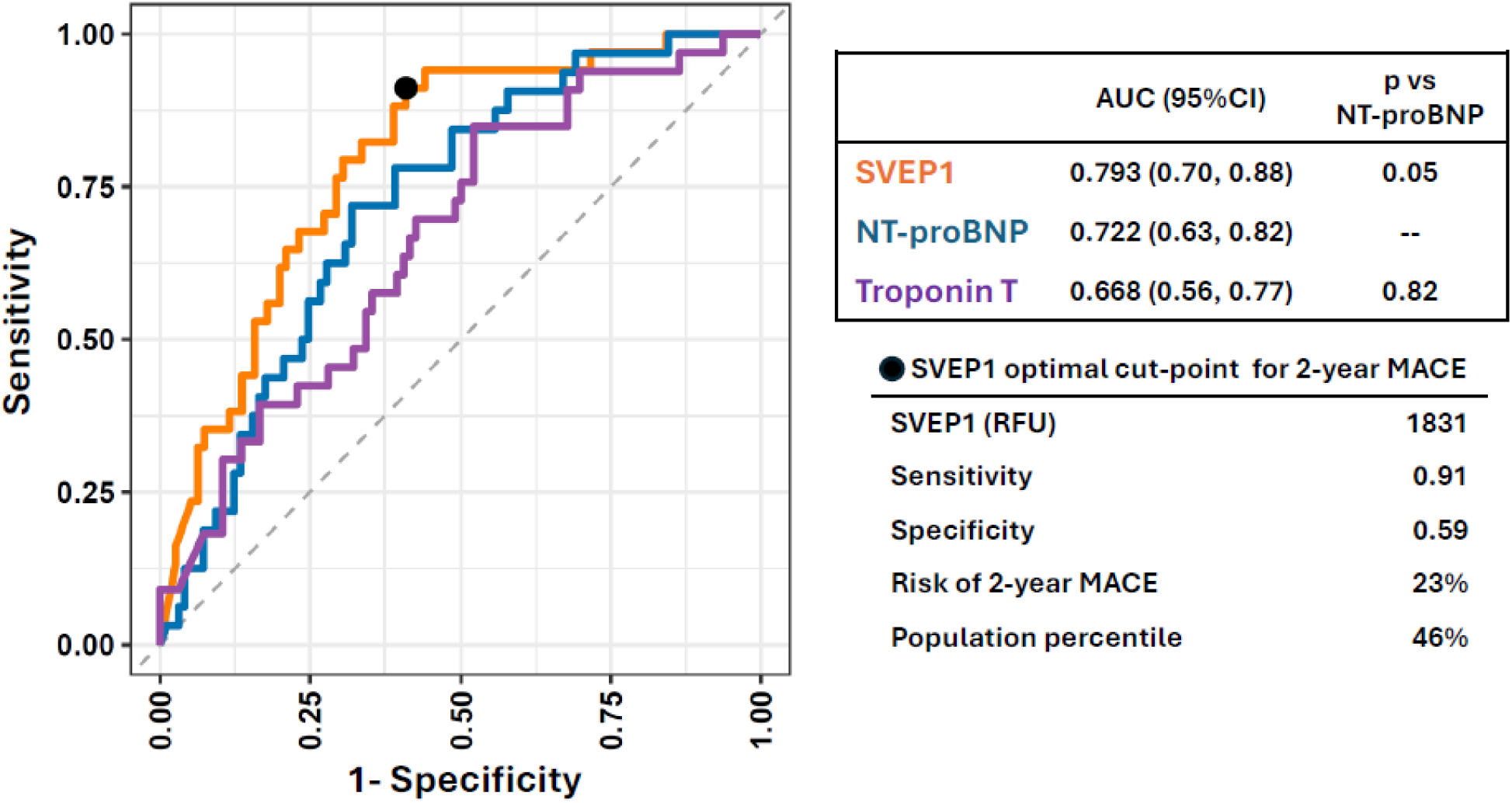
Receiver Operator Curves for Two-Year MACE. **Legend:** Receiver operator curves (ROC) for Sushi von Willebrand factor type A EGF and pentraxin domain-containing protein 1 (SVEP1), Troponin T, and N-terminal pro-B-type natriuretic peptide (NT-proBNP) for two-year MACE in the 20% test set of the pooled CRIC + PACE cohort (N=129), prevalent period. ⚫ = optimal cut-point for SVEP1 with the highest sensitivity and specificity for MACE, determined by Youden’s index.

### Analyses of SVEP1 For Specific CVD Outcomes, Subgroups and Alternate Time Horizons

Among the four individual outcomes within MACE, SVEP1 showed strongest associations for HF and CVD death. In the pooled cohort of CRIC and PACE (N=647), prevalent time point and using full follow-up, HR per log_2_ for MACE was 2.0 (p=3.2×10^-15^); for HF 2.0 (p=1.8×10^-9^); for CVD death 1.7 (p=5.2×10^-6^); for MI 1.3 (p=0.10); for stroke 1.6, p=0.06. **(Supplement 19).** Demographic subgroups were consistent in SVEP1 concentration and HR per log2 for MACE (p>0.1 for interaction) (**Table 4**). In CRIC, 5-year AUC(95%CI) was 0.696 (0.56, 0.82), and in PACE, 0.738 (0.61, 0.80). In CRIC, 10-year AUC(95%CI) was 0.738 (0.58, 0.83), and in PACE, 7-year AUC(95%CI) was 0.715 (0.59, 0.78). In PACE validation cohort, this 7-year AUC for SVEP1 surpassed the Expanded Refit PCE (0.587 (0.51, 0.66) (p<0.001). **(Supplement 16).**

**Table 4.** Subgroup Analysis for SVEP1 in the Pooled Cohort.

| Subgroup | SVEP1 |  |  |  |  |
| --- | --- | --- | --- | --- | --- |
|  | N patients<br>(N events) | Median [IQR]<br>(RFU) | HR per<br>log2 | p for HR |  |
| Incident time period | 1048<br>(480) | 3763<br>[2769, 5443] | 1.63 | 5.17 x10<sup>-13</sup> |  |
| Prevalent time period | 647 (317) | 3561<br>[2728, 5278] | 1.98 | 3.17 x10<sup>-15</sup> |  |
| Prevalent subgroups |  |  |  |  | p for<br>interaction |
| Age ≥ 60 years | 312 (188) | 3660<br>[2892, 6223] | 1.76 | 10.0 x10<sup>-7</sup> | 0.16 |
| Age < 60 years | 335 (129) | 3412<br>[2570, 4861] | 2.19 | 6.7 x10<sup>-9</sup> |  |
| Male | 383 (189) | 3527<br>[2690, 5205] | 2.08 | 1.7x10<sup>-10</sup> | 0.56 |
| Female | 264 (132) | 3597<br>[2777, 5667] | 1.89 | 2.0x10<sup>-6</sup> |  |
| Black race | 424 (207) | 3544<br>[2731, 5102] | 2.09 | 2.0x10<sup>-11</sup> | 0.84 |
| Non-Black race | 223 (110) | 3574<br>[2680, 5703] | 1.86 | 1.5x10<sup>-5</sup> |  |
| Diabetes | 452 (243) | 3580<br>[2731, 5495] | 1.91 | 1.1x10<sup>-10</sup> | 0.83 |
| No Diabetes | 195 (74) | 3527<br>[2651, 5212] | 2.17 | 9.5x10<sup>-6</sup> |  |
| CVD | 328 (204) | 3621<br>[2779, 5980] | 1.89 | 5.1x10<sup>-9</sup> | 0.63 |
| No CVD | 319 (113) | 3428<br>[2627, 5057] | 2.12 | 2.3 x10<sup>-7</sup> |  |
| >18 months on dialysis | 303 (160) | 3485<br>[2657, 5067] | 2.10 | 9.8x 10<sup>-10</sup> | 0.35 |
| ≤18 months on dialysis | 344 (157) | 3628<br>[2765,5453] | 1.90 | 2.5 x10<sup>-7</sup> |  |
| Unadjusted Cox HR per log2 for SVEP1 and MACE in the pooled CRIC and PACE cohort, using all follow-up for MACE. In the pooled cohort, incident time period denotes proteomics measured at median IQR 5.39 [3.02,9.08] months since dialysis initiation; prevalent time period refers to proteomics drawn at median IQR 18.4 [15.4,22.8] months since dialysis initiation. |  |  |  |  |  |

## DISCUSSION

We present, to our knowledge, the largest study of circulating proteomics and cardiovascular outcomes performed to date in patients with kidney failure undergoing hemodialysis, including 1048 study participants in two independent cohort studies sponsored by the National Institutes of Health(NIH) and incorporating a total of 10.7 million individual protein measures performed during incident and prevalent time periods relative to hemodialysis initiation. Twenty-two proteins reached Bonferroni significance of p<8×10^-6^ in CRIC over 14-year follow-up subsequent to Visit 2, when most participants had been on dialysis ≥ 1 year. Among these, proteins with successful external validation in PACE included SVEP1, Complement component C7, R-spondin 4, Tenascin, Fibulin-3 and Fibulin-5. Remarkably, over 2-year follow-up in CRIC, SVEP1 as a single protein predicted the composite CVD outcome better than a model based on multiple conventional risk factors including TnT and NT-proBNP.

SVEP1 emerged as a statistically significant predictor of MACE with Bonferroni correction for ∼ 7000 tests (i.e., p<8×10^-6^) when measured at CRIC Visit 1 (HR per log_2_ 1.6, p=2.9×10^-8^), or Visit 2 (HR per log_2_ 1.8, p=2.1×10^-12^), or as change in SVEP1 occurring between the two CRIC proteomic time points (HR per 25% increase 1.1, p=6.8×10^-6^. **(Supplements 5,7,12).** In PACE, SVEP1 at time point 1 or 2 met FDR<0.05 for validation, and percent change in SVEP1 was nominally significant (p=0.01) **(Supplements 11,13).** In alignment with data collected by the United States Renal Data System(USRDS),^27^ 10% of participants experienced MACE within two years following biomarker measurement. The importance of near-term cardiovascular risk prediction in populations with high events rates is supported by the National Heart, Lung, and Blood Institute^28^, and provides a strong rationale for focusing on 2-year risk models. For predictive risk modeling in CRIC, the gain in AUC for SVEP1 was most significant for short-term (2-year) prediction (**Figure 3**), while in PACE validation cohort SVEP1 surpassed the Expanded Refit PCE that included TnT and NT-proBNP, for events within 7 years (**Figure 4**).

Originally named polydom, SVEP1 is a 390 kDa protein that consists of many domains, including 34 complement control modules, or Sushi domains, structural motifs known for complement regulation.^29^ It also has a pentraxin domain, which typically activates complement,^30^ Von Willebrand factor A domain, ephrin receptor-like domain, and several epidermal growth factor-like domains.^31,32^ SVEP1 is produced by activated fibroblasts,^32^ adipocytes^33^ and vascular smooth muscle cells.^26^ SVEP1 has numerous receptors, including integrins with roles in hypertension^34^ (α9β1^35^) and vasoconstriction (α4β1)^36^. Platelet and endothelial aggregation receptor-1 (PEAR1) is a receptor for SVEP1 that activates target of rapamycin(mTOR).^32^ Mendelian randomization and animal studies provide strong evidence that SVEP1 is atherogenic^26^ and directly activates platelets.^32^ Circulating SVEP1 has now been measured by aptamers (SomaScan) in several non-dialysis settings, showing strong associations with diseases of vascular etiology, including MI,^26^ and dementia.^37^ We have previously observed a striking association of SVEP1 with incident HF in 2906 CRIC participants with CKD stages 3 and 4: after adjustment for estimated glomerular filtration rate, SVEP1 predicted incident HF at p<10^-30^ (unadjusted p<10^-40^). ^8^ Circulating SVEP1 has been shown to predict HF in non-CKD populations^38,39^ and death in HF populations.^40,41^ The present study is the first to show a strong association of SVEP1 with MACE, and particularly with HF and cardiovascular mortality in patients with kidney failure on hemodialysis. Especially striking is the finding that the strongest model of MACE created with elastic net regression consisted of SVEP1 alone as a single protein, a unique finding given that proteomic risk models derived by applying elastic net to proteomic datasets typically consist of tens or even hundreds^42^ of proteins.

Twenty-two proteins met a statistical threshold of p<8×10^-6^ in CRIC at Visit 2 (prevalent time period) **(Supplement 7).** With the exception of brain natriuretic peptide (HR per log_2_ 1.5, p=4.4×10^-11^), these top proteins are not commonly considered as CVD biomarkers among patients undergoing hemodialysis. Among these 22 proteins, we highlight 12 in **Table 3** that passed external validation in PACE. All twelve retain significance (p<0.05) after adjusting for clinical risk factors and accounting for competing risk. Spline analyses shown in **Figure 1** demonstrate predominantly linear associations within the interquartile range, as opposed to U-shaped associations observed for traditional risk factors such as cholesterol^43,44^ in the dialysis population. Replication, independence and linearity are favorable qualities for biomarkers. Moreover, proteins such as SVEP1 (390kDa), Complement component 7(94kDa) and Tenascin(180kDa) are too large to be cleared renally or by hemodialysis. If developed further as CVD biomarkers for patients with advanced CKD or kidney failure, their larger size might be an advantage over smaller sized troponins and natriuretic peptides, whose circulating levels are affected by reduced renal clearance. Our analyses of change in proteins suggest that SVEP1, R-spondin 4 and Tenascin are dynamic risk factors. As compared to static factors such as age, sex, or most comorbid conditions, proteins are more likely to vary in response to patient’s diet,^45^ medications,^46^ or potentially with adherence to dialysis. It is conceivable that a biomarker panel could be developed as a modifiable risk stratification tool.

Complement components and complement-related proteins had the highest statistical significance in functional enrichment at p<10^-17^ **(Supplement 15).** Patients with kidney failure on hemodialysis or peritoneal dialysis experience chronically high complement activation due to bioincompatibility of dialysis membranes, peritoneal dialysis fluids, and frequent infectious exposures.^47,48^ Genetic studies of complement inhibitor factor H^49^ and complement receptor 1^50^ support causal links with CVD in kidney failure, similarly to non-CKD populations.^51,52^ Sushi proteins such as SVEP1 have structural motifs called complement control protein (CCP) modules, known for complement interactions.^29,31,53^ SVEP1 has 34 CCP, as well as a pentraxin domain (that typically activates complement)^30^. It is likely that SVEP1 plays a role in complement regulation/activation, a hypothesis worthy of future investigations. Besides complement pathways, the growth hormone(GH) / insulin growth factor (IGF) axis featured prominently in our network analyses (**Figure 2**). The diverse effects of the GH/IGF1 axis on CVD have been studied extensively.^54^

Our study has many strengths. CRIC and PACE are prospective observational studies funded by the NIH with excellent phenotyping of the participants initiated on hemodialysis, exemplary blood sample collection, and meticulous adjudication of CVD outcomes. The heterogeneity between CRIC and PACE enhances the generalizability of the results. Our previously published quality control studies of SomaScan^16^ and external validation for top proteins in PACE support the rigor of our findings. A unique perspective is gained by fitting the conventional risk factor model (Expanded Refit PCE) and novel protein(SVEP1) to the same cohort, facilitating a head-to-head comparison of conventional and protein models.

We also acknowledge limitations. High event rates contributed statistical power to the analyses, but the relatively modest sample sizes limited the number of sub-group analyses, an issue we addressed by pooling the two cohorts. The effect of the hemodialysis procedure itself on the circulating proteome, and comparisons to patients on peritoneal dialysis were beyond the scope of this manuscript and should be the subject of future studies.

In conclusion, by using large-scale proteomics we have identified novel biomarkers and potential mediators of CVD in patients undergoing maintenance hemodialysis. Among 6287 proteins, SVEP1 emerged as the strongest predictor of the MACE outcome. Future studies could elucidate the extent to which predictive protein biomarkers have causal roles in CVD and whether they can be modified by pharmacological or lifestyle interventions, or by dialysis.

## Data Availability

The CRIC data included in these analyses will be available to approved requestors in the future in the NIDDK Central Repository and at dbGaP. Prior to the availability of data in the repositories, requests can be made to the CRIC Study group by contacting the CRIC Scientific and Data Coordinating Center at. Appropriate regulatory and scientific approvals are required. The data are not publicly available due to individual-level informed consent restrictions.

## Non-standard Abbreviations and Acronyms

MAD: median absolute deviation
SVEP1: Sushi von Willebrand factor type A EGF and pentraxin domain-containing protein 1
ANML: Adaptive Normalization by Maximum Likelihood
IGFBP: Insulin growth factor binding proteins

## Acknowledgements

*CRIC Study Investigators: Lawrence J. Appel, MD, MPH; Debbie L Cohen, MD; Laura M Dember, MD; Alan S. Go, MD; James P. Lash, MD; Mahboob Rahman, MD; Vallabh O. Shah, PhD, MS; Mark L. Unruh, MD, MS

## Data Sharing Statement

A portion of the data reported here have been supplied by the United States Renal Data System (USRDS). The interpretation and reporting of these data are the responsibility of the author(s) and in no way should be seen as an official policy or interpretation of the U.S. government.

