## Supplemental Methods for "Discovery and Validation of SVEP1 and Other Novel Cardiovascular Biomarkers For Patients with Kidney Failure On Maintenance Hemodialysis"

***Participants***

The Chronic Renal Insufficiency Cohort(CRIC) was designed to investigate risk factors for progression of CKD, incident CVD, and overall mortality in persons with CKD.^1^ Between 2003 and 2008, CRIC enrolled a total of 3,939 ethnically diverse men and women at 13 sites affiliated with 7 clinical centers, ages 21–74 years, with estimated glomerular filtration rate 20–70 ml/min/1.73m^2^ by the simplified (4-variable) Modification of Diet in Renal Disease equation.^1^ Eligibility criteria and baseline characteristics of the CRIC cohort have been published.^1,2^ The CRIC study was approved by the Institutional Review Boards of the participating centers, and the research was conducted in accordance with the principles of the Declaration of Helsinki. All study participants provided written informed consent. Between 2004 and 2020, 1123 CRIC participants reached kidney failure and started dialysis. Among these participants, 251 (22.4%) had no further study visits and another 232 (20.7%) provided no blood samples after starting dialysis. Due to the potential interference of lupus antibodies with aptamers (communication from SomaLogic), we excluded 10 (0.9%) CRIC participants with systemic lupus erythematosus. For the present analysis, we included 630 CRIC participants with kidney failure receiving maintenance dialysis, enrolled from all 13 sites at 16 different study visits **(Supplemental Table 1).** We ran SomaScan V4.1 on plasma EDTA samples drawn at the first study visit following initiation of dialysis; we assayed samples on 421 (66.8%) participants who remained on hemodialysis a mean of 1.1 ± 0.8 years following the first proteomics sample. Based on *post-hoc* interviews with CRIC study sites, approximately 80% of blood samples were drawn on a non-dialysis day, with the remainder drawn on the dialysis day prior to initiation of the hemodialysis procedure. Seventy-five percent of CRIC participants were fasting at the time of blood draw. Blood was promptly centrifuged, and plasma was sent on dry ice to the Central Lab at University of Pennsylvania where it was aliquoted and stored at -80^o^C.

The Predictors of Arrhythmic and Cardiovascular Risk in ESRD(PACE) study was designed to evaluate risk factors for sudden cardiac death in patients with kidney failure who had recently been started on hemodialysis. PACE enrolled participants from 27 dialysis units in the Baltimore area; there were four annual study visits over the course of 2008–2012. Participants were ≥18 years old, English-speaking, and had initiated thrice weekly in-center hemodialysis fewer than six months earlier. Exclusion criteria included cancer, presence of pacemaker or automatic implantable cardioverter defibrillator, and other criteria previously described.^3^ Information on glomerulonephritis, but not specifically lupus, was collected in PACE. We assayed SomaScan 4.1 on plasma EDTA samples from 418 participants at Visit 1 and 226 participants from Visit 2 (separated by mean (SD) 1.1 (0.2) years). PACE fasted blood samples were collected on a non-dialysis day. Plasma tubes were centrifuged immediately and stored in a -20^o^C freezer; within 2–4 hours, samples were aliquoted and transferred to a -80^o^C freezer.

***SomaScan Assay and Quality Control***

SomaScan is an assay based on modified aptamers, which are chemically modified single strands of deoxyribonucleic acid ~40 nucleotides long, as binding reagents for target proteins.^4-9^ Modified aptamers bind to proteins with high affinity similar to antibodies (lower limit of detection 10^-15^ moles per liter.)^4,6,7^ “Pull-down” studies, in which the aptamer-protein complexes were isolated and the identities of the bound proteins were verified by targeted mass spectrometry and gel electrophoresis, have been performed for 920 proteins among 1305 proteins in a previous version of the assay.^8^ These studies showed that >95% of aptamers correctly targeted the intended proteins (for those proteins in concentrations sufficient to be detected by mass spectrometry). Samples on the SomaScan assay are run at three different dilutions to assay each analyte within its linear range of concentrations. Assay results are quantified on a hybridization microarray and reported in relative fluorescent units (RFU). SomaLogic has procedures for data calibration, standardization and internal controls, typical of microarray technologies. SomaLogic normalizes the entire protein dataset using Adaptive Normalization by Maximum Likelihood (ANML) to remove unwanted biases in the assay. ANML is an iterative procedure that adjusts values for analytes that fall outside expected measurements from a reference distribution.^10^

The SomaScan V4.1 menu includes 7596 aptamers. We excluded 308 aptamers paired with non-human proteins, and 117 incompletely characterized investigational aptamers. Circulating concentrations of numerous analytes are 2- to 10-fold higher in patients with kidney failure compared to those without kidney disease. We excluded about 0.4% of aptamers (32 for CRIC, 31 for PACE) that saturated the SomaScan assay due to high concentrations. We included split duplicates of CRIC samples in CRIC batches to allow for quality control of assay precision. There were four proteins with intra-assay CV>50% in these duplicates, and these were excluded from CRIC and PACE analyses. Given that some proteins are measured by two or more aptamers, we analyzed 7135 aptamers (6287 unique proteins) in CRIC, and 7140 aptamers (6291 unique proteins) in PACE. Proteins included in the SomaScan V4.1 menu, and the reasons we excluded any protein, are listed in **Supplemental Table 2.** SomaScan intra-assay coefficients of variation (CV) from plasma of healthy individuals are reported as ≤5%.^11,12^ We previously published a quality control study of proteins in the SomaScan 4.0 platform^10^ (4607 unique proteins) in plasma samples of 40 participants on maintenance hemodialysis of the Cardiac, Endothelial Function and Arterial Stiffness in End-Stage Renal Disease (CERES) study. Median [IQR] intra-assay CV was 2.4% [1.8%, 3.4%]. Median[IQR] inter-assay CV was evaluated by inserting CERES blinded, duplicate samples in batches of CRIC and PACE samples run approximately one month apart, and was 7.4% [4.6%, 13.1%]. In CERES samples, short-term within-subject CV over one week was 5.8% [3.4%, 9.7%]. Also using CERES data, we explored the Adaptive Normalization by Maximum Likelihood(ANML) data format versus raw protein data for 1) assay variability and 2) protein fold change in patients who died versus survived over 2.5 years. Technical and short-term biological variability in paired samples were lower when we used ANML-formatted data, but fold-change in survivors was minimally affected by ANML format.^10^ We chose to use ANML formatted data for the current analyses in CRIC and PACE to facilitate comparison to other studies. In the present study we also analyzed intra- and inter-assay CVs for the 2298 proteins in the Somascan 4.1 that were not in version 4.0: median intra-assay CV was 3.3% and inter-assay CV 7.9% **(Supplemental Table 3)**.

The two different aptamers in SomaScan (SeqId11109-56 and SeqId 11178-21) target different regions of the SVEP1 protein, and binding of both aptamers to SVEP1 has been validated with mass spectrometry.^13^ In our dataset, the two SVEP1 aptamers correlated at *rho*=0.97 and had nearly identical HRs for the MACE outcome. (Correlation: **Supplemental Figure 1**; **Supplement 5-10**)

***Study Outcomes and Censoring***

The composite CVD outcome of major adverse cardiovascular events(MACE) consisted of time to myocardial infarction(MI), heart failure (HF), stroke, CVD death, or death of unknown cause. Deaths adjudicated as non-CVD were excluded from the composite outcome. In CRIC, cardiovascular events were adjudicated by two physicians after reviewing imaging, laboratory data, hospital notes and death certificates, and categorized as probable or definite MI, HF, stroke or CVD death, as per published protocols.^1,14^ We included probable and definite events in these analyses. Deaths were ascertained from next of kin, death certificates, obituaries, hospital records, the Social Security Death Master File, and the National Death Index. Since not all deaths occurred in a hospital, some could not be adjudicated by review of hospital reports. The CRIC Statistical Data Coordinating Center (SDCC) biostatisticians designed a machine learning algorithm to determine if a death was cardiovascular-related based on causes of death listed on the death certificate. There was a small proportion of deaths labelled as “unknown cause” and given the frequency of sudden cardiac arrests in this population,^15^ we included these as CVD deaths in the composite outcome of MACE. Outcomes for this analysis were adjudicated through May 2020.^16^ In our analyses, survival time was censored for kidney transplantation (N=88 participants for time starting from first visit). Participants were queried about dialysis modality at each study visit, and those who had changed to peritoneal dialysis were not included in the samples from the first or second time point. However, dialysis modality was not specifically documented on the dates of clinical outcomes.

In PACE, events were adjudicated by two independent reviewers on the PACE Endpoint Committee, and any discrepancies were resolved by a third reviewer. Criteria for non-fatal MI, HF, stroke were similar to CRIC and have been published.^3^ Mortality was ascertained using reports from dialysis units confirmed with Centers for Medicare and Medicaid Services Form 2746. CVD mortality was defined as death arising from arrhythmia, ischemic CVD, or ischemic [cerebrovascular disease](https://www.sciencedirect.com/topics/medicine-and-dentistry/cerebrovascular-disease), and [sudden cardiac death](https://www.sciencedirect.com/topics/medicine-and-dentistry/sudden-cardiac-death). Sudden cardiac death was defined as a sudden pulseless condition (collapse or syncope) presumed to be due to an arrhythmia occurring out of the hospital or in the emergency room in an otherwise stable individual. If the event was not witnessed, there had to be evidence that the patient was seen in a stable condition within the 24 hours preceding the event or since the last dialysis session. None of the events that occurred during a hospitalization, in a nursing home or hospice were classified as sudden cardiac death. Deaths were attributed to coronary artery disease if the patients (1) had a definite MI within 4 weeks of death, or (2) had chest pain within 72 hours of death in cases of out-of-hospital death or cardiac pain in cases of in-hospital death, or (3) history of chronic ischemic heart disease such as MI, coronary insufficiency, or angina pectoris, or (4) the underlying cause of death in the death certificate included ICD-10 code I20, I21, I22, I23, I24, I25, I46, I51.6, I51.9, R99, J96, if there was no evidence of a non-atherosclerotic or non-cardiac atherosclerotic process that was the probable cause of death.^3^ Outcomes for the current study were adjudicated through December 2015. In our analyses, survival time was censored at kidney transplantation (N=59 for time starting from first visit) or change to peritoneal dialysis (N=17 during time starting from first visit). Outcome and censored events are tabulated in **Supplemental Table 1.**

***Covariate Definitions***

In CRIC, sociodemographic data were obtained at baseline using self-reported questionnaires, including sex, self-reported race/ethnicity, and smoking status. Self-reported comorbidity data were updated at each study visit. Diabetes mellitus was defined by fasting glucose ≥126 mg/dL and/or the use of insulin or oral hypoglycemic medications. Hypertension was defined by a systolic blood pressure(SBP) ≥140 mm Hg, diastolic blood pressure(DBP) ≥90 mm Hg, or the use of antihypertensive medications. Prevalent or new onset CVD was assessed at each study visit (including study visits for patients who had initiated dialysis) by a self-reported history of prior MI, coronary revascularization, heart failure, stroke, or peripheral artery disease. Body mass index(BMI) was calculated using measured height and weight and expressed in kilograms per meter squared. Serum lipids and phosphate were measured at study baseline (concurrent with proteomic assays during the incident time period), at CRIC Central Lab specifically for the current study. In PACE, data for these covariates were collected in manners similar to CRIC and are described in the Study Design.^3^

***Statistical Analysis***

*Baseline characteristics and protein normalization*

Continuous values of participants’ baseline characteristics were summarized as median[IQR]. We utilized SomaScan protein values measured in relative fluorescent units that had been normalized using ANML, with subsequent median absolute deviation (MAD) based standardization and Winsorizing (outlier clipping) at the median ± 5 MAD, as we have previously published.^17^ For a data set, $x_{1}, x_{2}, x_{3}, \ldots, x_{n}$, the MAD is defined as the median of the absolute deviation from the data’s median: MAD = median(|$x_{i}$ - $x_{median}$|), where $i$= $1,2,3,\ldots,n$, and $x_{median}$ is the median of the data set. For each protein, we first capped the extreme values by +/-5*MAD and then standardized it by subtracting the median and dividing by the MAD.

*Imputation for missing clinical or laboratory variables*

We performed multiple imputation in most instances of missing variables required for the analyses, which are listed herein as % missing at incident time point, % missing at prevalent time point. We applied multiple imputation using chained equations as implemented in the R package *mice*. Five imputed datasets were created with synthesized estimates and attendant standard errors obtained via Rubin’s pooling method. For CRIC, multiply imputed variables were SBP (5%, 6%), DBP (5%, 6%), hemoglobin (6%, 1%), phosphate (0.3%, 80%), calcium (4%, 0%) and albumin (4%, 0%). PACE multiply imputed variables were SBP (9%, 10%), DBP (9%, 10%), BMI (0.5%, 12%), as well as laboratory markers that were missing in 12% of patients at the prevalent time point in PACE (serum parathyroid hormone (PTH), phosphate, hemoglobin, calcium, albumin). In CRIC, PTH (80%, 85%) was based on a regression model developed in CRIC participants with both aptamer and traditional PTH assay. (CRIC *rho* for PTH = 0.83). Lipids (low-density lipoprotein cholesterol (LDL-C), high-density lipoprotein cholesterol (HDL-C), triglycerides, total cholesterol) were missing at second time points for both cohorts: CRIC (1%, 100%) and PACE (9%, 100%). We performed elastic net regression on all SomaScan proteins separately in CRIC and PACE for samples with aptamers and measured lipids and then utilized the elastic net model to calculate imputed lipid values where missing in the respective cohorts. Scatterplots for aptamer-based values and traditional assay values of PTH and lipids are shown in **Supplemental Figure 2**. Hybrid clinical-protein models were developed using elastic net, which is not amenable to multiply imputed variables. In the pooled cohort we ran elastic net in a dataset with proteins, Pooled Cohort Equation (PCE) variables, and dialysis-related factors: (log(DBP), log(BMI), log(triglycerides), serum albumin, hemoglobin, log(time since dialysis initiation), log(calcium), log(phosphate), log(PTH)); analyses were done for the 2, 5- and 10-year time horizon. Due to high missingness in some of the variables, we used the average of 5 values for each imputed variable. Since no dialysis factors were selected, we did not fit additional models with dialysis-related factors. Hybrid protein-PCE models were developed on datasets utilizing the median value at corresponding visit for SBP (5% missing) and modal values for hypertension treatment (6% missing).

*Single protein associations with the composite cardiovascular outcome (MACE)*

The primary outcome for the study was the composite of non-fatal MI, HF, stroke, and CVD death or death of unknown cause (MACE). We used Cox proportional hazards regression to assess the association between individual proteins and time to event. We performed an initial survey of single protein associations normalized to MAD; this approach allows for the ranking of predictors and is more robust for skewed data than conventional methods of standardization (mean subtraction, standard deviation division). We focused on ‘top hits’ from among the protein associations (HR significant in CRIC at Bonferroni correction of p<8x10-6). ^18,19^ Top hits are presented as HR per log_2_, a more widely used standardization that facilitates interpretation of effect size and comparison to other studies. Since bidirectional or U-shaped associations of traditional risk factors such as cholesterol in the hemodialysis population are less useful for risk stratification,^20^ for we examined top proteins using restricted cubic splines with 4 knots at 5th, 35th, 65th, 95^th^ percentiles. To account for the high rates of non-CVD death in CRIC and PACE, which mirrored data in the United States Renal Data System,^21^ we adjusted for competing risks by designating non-CVD death as a competing event and implementing the Fine Gray method.^22^ In order to gauge independence of novel biomarkers and traditional risk factors, we created an adjusted model that included PCE variables: age, sex, self-reported race, diabetes, SBP, treatment for hypertension, smoking, HDL, and total cholesterol. We then created a fully adjusted model with PCE variables and potential confounding risk factors for patients undergoing hemodialysis, prioritizing factors associated with the composite outcome in CRIC at p<0.05 (DBP, BMI, triglycerides, albumin) or PACE (hemoglobin), and including additional plausible confounders without significant associations in these cohorts (time since dialysis initiation and serum calcium, phosphate, PTH).The fully adjusted model included the following variables, with missing values imputed by multiple imputation, or for lipids, and PTH, using aptamer-based equations as described above: age, designated race, sex, log(total cholesterol), log(HDL), SBP, hypertension treatment, diabetes, current smoking; Dialysis-related factors: log(DBP), log(BMI), log(triglycerides), serum albumin, hemoglobin, log(time since dialysis initiation), log(calcium), log(phosphate), log(PTH).

*Network analysis and functional enrichment*

To better understand biological pathways represented by our ‘top hit’ proteins, we analyzed the subset of proteins significant at FDR<0.01 (corresponding to p<1x10-4) in CRIC unadjusted analyses of time point 2. This statistical threshold was chosen for convenience, as it identified a manageable dataset for Search Tool for the Retrieval of Interacting Genes/Protein (STRING). Starting with 62 proteins, we utilized the Search Tool for the Retrieval of Interacting Genes/Proteins*(*STRING) interaction database using stringApp^23^ or Cytoscape^24^ and used an interaction score cutoff of 0.3, allowing 15 indirectly connected neighboring proteins to reconstruct a STRING network.^25^ We then used the largest network of 64 proteins to perform enrichment analysis, to investigate functional consistency among networked subsets of proteins. First, we performed over-representation analyses(ORA) using the databases of Gene Ontology,^26^ Wikipathways^27^ and KEGG.^28^ ORA is used to determine the proportion of proteins within a particular gene set that are found among the group of 64 proteins, and compare this proportion to the proportion that would be found using the background of all proteins measured by SomaScan. For a given protein that was measured by two or more aptamers, we employed the aptamer with the largest effect size. For a given aptamer that was annotated by multiple UniProt identifiers, we used the first identifier.^29-31^ Enriched gene sets that passed the FDR threshold of 0.05 were selected and redundant gene sets were filtered using the redundancy cutoff of 0.5 in StringApp.^23^

*Conventional Risk Models*

We aimed to evaluate novel protein CVD risk factors in the context of conventional CVD risk factors in the published PCE,^32^ as well as two established biomarkers in the kidney failure population, Troponin T and NT-proBNP.^33^ Three conventional models are used as comparators. The Original PCE includes published coefficients evaluated separately in Blacks and non-Blacks, and in men and women, using age, age x total cholesterol, age x HDL, age x treated SBP, age x smoking, diabetes, SBP, treatment for hypertension, smoking, HDL, total cholesterol. In an alternate version, we refit the PCE variables listed above plus sex and designated race, to the CRIC or pooled cohort 80% training set and evaluated the AUC(95%CI) in the 20% set of the respective datasets. The third conventional risk model, the Expanded Refit PCE, was devised by refitting PCE variables, plus aptamer measures of Troponin T and NT-proBNP to the 80% training set and testing discrimination in the 20% testing set. We previously have shown good correlation between traditional and aptamer assays for Troponin T (*rho* 0.68) and NT-proBNP(*rho* 0.80) in plasma of patients with kidney failure,^10^ and for the Expanded Refit Pooled Cohort Equation + Troponin T and NT-proBNP model, we utilized aptamer values for these two markers.

*Protein Risk Models for MACE*

Aiming to develop multi-protein and hybrid clinical-protein models, we ran elastic net regression in three different sets of candidate predictors: 1) 6287 proteins in Somascan; 2) 6287 proteins + PCE variables; 3) in the pooled cohort, we evaluated 6287 proteins, PCE variables, and dialysis-related factors (time since dialysis initiation, DBP, BMI, triglycerides, albumin, hemoglobin, calcium, phosphorus, parathyroid hormone). For several scenarios, the elastic net model chose only proteins and no clinical factors, resulting in no clinical-protein hybrid model. Risk models were derived in a training set obtained as a random partition of 80% of the CRIC participants, tested in the remaining 20%, then validated in the full PACE cohort. In the pooled analysis of CRIC and PACE, each conventional or protein model was trained and then tested in 80% and 20% sets, respectively. For risk models developed using elastic net in the pooled cohort, we considered an indicator variable to account for any differences in the CRIC or PACE cohorts. The indicator variable was not selected by elastic net when allowed to compete with other variables; forcing in the indicator variable resulted in a modest increase or decrease in AUC for some models. We show models with the indicator variable in **Figure 3** and alternate models in **Supplement 18**. Our frontline technique for developing protein risk prediction models was elastic-net Cox regression which combines ridge (L2) and Least Absolute Shrinkage and Selection Operator (L1) penalties. The relative contributions of the two penalties are controlled by a mixing parameter 𝛼 which we set to 0.5 for balance. The shrinkage (regularization) parameter 𝜆 which controls model complexity (the number of included proteins) was determined by 10-fold cross validation and the “1 standard error rule” (for parsimony). After the final selection of proteins, to reduce bias in estimated regression coefficients,^34^ we refit the selected features for the elastic-net model in another Cox regression, as previously published.^14^ AUCs for risk models were compared using paired bootstrapped t-tests with one-sided p-values,^35^ and p <0.05 was deemed statistically significant. Due to the small sample available in the testing sets, we assessed calibration for refit models in the training set using a model-based test that can accommodate survival endpoints in addition to continuous and binary outcomes.^36^ Calibration for the Original PCE was assessed using Greenwood-Nam-D'Agostino test.^37^

We compared SVEP1 as a single biomarker to NT-proBNP and Troponin T by evaluating and comparing AUCs built from risk models trained in the 80% set of the pooled cohort, and tested in the 20% set. We formally compared single protein risk models with paired bootstrapped t-tests and one-sided p-values, as above. We evaluated the optimal cut-point with highest sensitivity and specificity for SVEP1 by selecting the point with the highest value Youden statistic.

*External Validation*

We validated in PACE, full cohort and full follow-up time, the 22 proteins associated with MACE outcome at p<8x10-6 in CRIC, designating FDR<0.05 as criterion for replication. Risk models, either with published coefficients (as for PCE) or with coefficients fit to the CRIC 80% testing set, were also validated in PACE, full cohort. We conducted statistical analyses using R, version 4.5.0 (RStudio, Inc., Boston, MA. URL <http://www.rstudio.com/>), with the packages of *glmnet* (version 4.1.8), *survival* (version 3.8.3), *survivalROC* (version 1.0.3.1), *survcomp* (version 1.58.0), *tidyverse* (version 2.0.0), *magrittr* (version 2.0.3), *mice* (version 3.17.0), *flextable* (version 0.9.7), *officer* (version 0.6.8) *harmonicmeanp* (version 3.0.1), and *plotRCS* (version 0.1.5).
